# SoK: Intelligent Detection for Polycystic Ovary Syndrome(PCOS)

**DOI:** 10.1101/2024.12.25.24319623

**Authors:** Meng Li, Zanxiang He, Liming Nie, Liyun Shi, Mengyuan Lin, Minge Li, Yanjun Cheng, Hongwei Liu, Lei Xue

## Abstract

Graphical Abstract

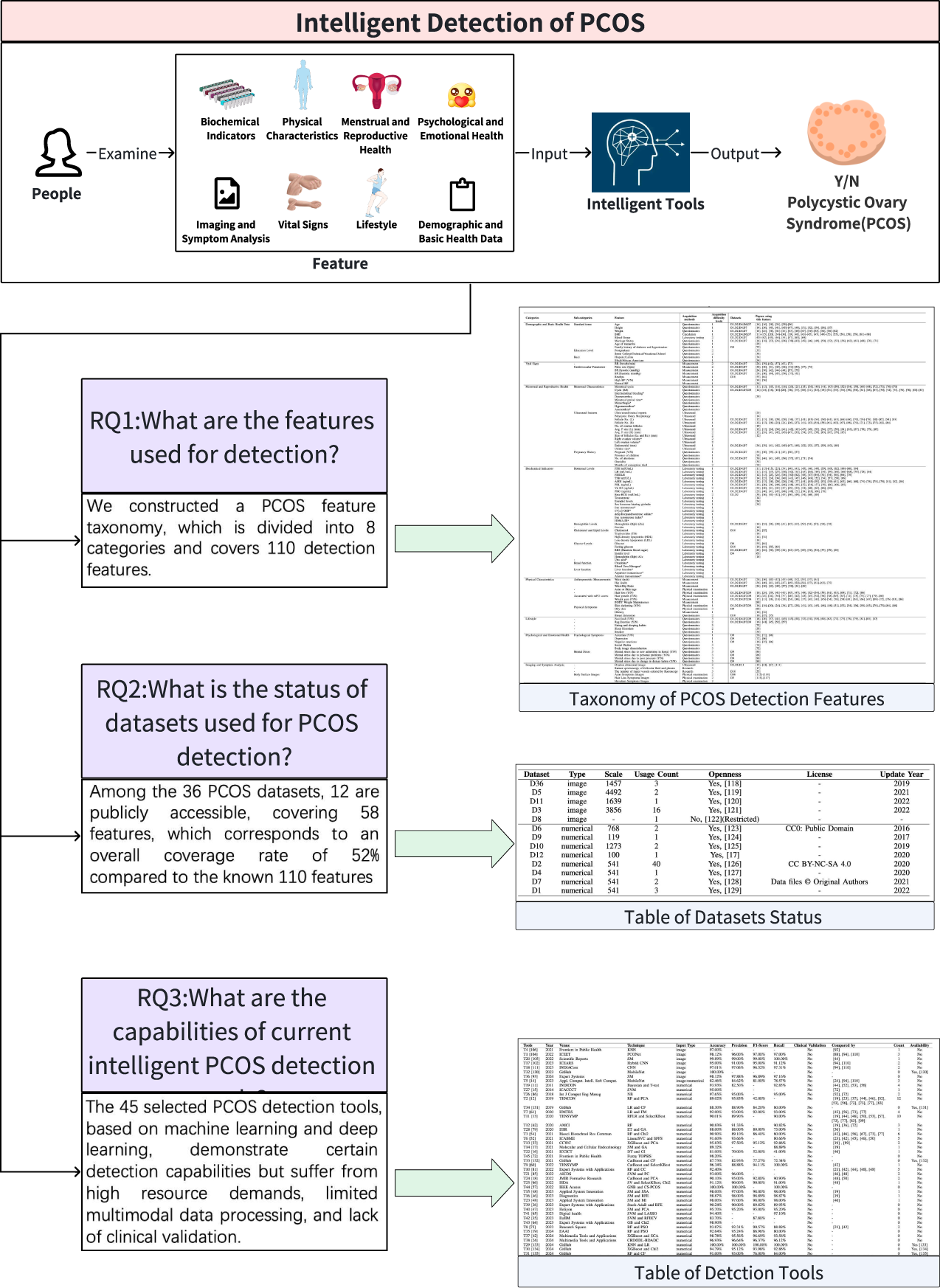

**Highlights:**

- Conducted a systematic review of the existing literature, focusing on Polycystic Ovary Syndrome intelligent detection, and constructed the comprehensive taxonomy for PCOS detection features to date, providing a standardized reference for future research.
- Systematically evaluated the capabilities and limitations of current intelligent PCOS detection tools, offering valuable guidance for the development of more efficient and accurate tools.
- Thoroughly analyzed the current status of 12 publicly available datasets used for PCOS detection, providing clear directions for future dataset development in this field.
- Made the analysis results publicly available, providing data resources and references for researchers, with the aim of advancing the field of intelligent PCOS detection.

Recent research in the field of Polycystic Ovary Syndrome (PCOS) detection has increasingly utilized intelligent algorithms for automated diagnosis. These intelligent PCOS detection methods can assist doctors in diagnosing patients earlier and more efficiently, thereby improving the accuracy of diagnosis. However, there are notable barriers in the field of intelligent PCOS detection, including the lack of a standardized taxonomy for features, inadequate research on the current status of available datasets, and insufficient understanding of the capabilities of existing intelligent detection tools. To overcome these barriers, we propose for the first time an analytical framework for the current status of PCOS diagnostic research and construct a comprehensive taxonomy of detection features, encompassing 110 features across eight categories. This taxonomy has been recognized by industry experts. Based on this taxonomy, we analyze the capabilities of current intelligent detection tools and assess the status of available datasets. The results indicate that 12 publicly available datasets, the overall coverage rate is only 52% compared to the known 110 features, with a lack of multimodal datasets, outdated updates and unclear license information. These issues directly impact the detection capabilities of the tools. Furthermore, among the 45 detection tools require substantial computational resources, lack multimodal data processing capabilities, and have not undergone clinical validation. Based on these findings, we highlight future challenges in this domain. This study provides critical insights and directions for PCOS intelligent detection field.

## 1. Introduction

Polycystic Ovary Syndrome (PCOS) is one of the most common gynecological endocrine disorders. Its clinical characteristics primarily include clinical or biochemical hyperandrogenism, chronic anovulation, and polycystic ovarian morphology, often accompanied by insulin resistance and obesity [1, 2]. In recent years, the prevalence of PCOS has been increasing. Globally, using the Rotterdam criteria, the prevalence of PCOS is estimated to be between 10% and 13%, significantly affecting women’s quality of life and becoming a major public health issue worldwide [3]. Moreover, PCOS is associated with metabolic syndrome, type 2 diabetes, cardiovascular disease, psychological disorders, and other metabolic disturbances. These issues highlight the critical need for early diagnosis and long-term management [4, 5, 6, 7].

Currently, the most widely adopted diagnostic criteria for PCOS are the Rotterdam criteria. These define PCOS as the presence of two out of the following three conditions, excluding other causes of hyperandrogenism: oligoor anovulation, clinical or biochemical signs of hyperandrogenism, and polycystic ovarian morphology [1, 8, 9]. However, traditional medical approaches to diagnosing PCOS still face significant challenges. They are often subjective, heavily reliant on clinicians’ expertise, and can be invasive and uncomfortable for patients. Additionally, the heterogeneity and individual variability of symptoms may result in misdiagnosis or delayed diagnosis [10].

To address these challenges, researchers have increasingly explored the use of intelligent technologies, including machine learning and deep learning, to enhance the accuracy and efficiency of PCOS diagnosis. Various methods have emerged to detect PCOS using intelligent techniques. For instance, in 2011, Mehrotra et al. [11] proposed an automated PCOS detection method based on a Bayesian classifier utilizing clinical and metabolic parameters. In 2019, Denny et al. [12] developed a system using Random Forest Classifier with minimal yet promising clinical and metabolic parameters for early detection and prediction of PCOS. Similarly, in 2020, Bharati et al. [13] presented a tool based on hybrid Random Forest and Logistic Regression (RFLR) for reliable classification of PCOS patients. Most recently, in 2023, Alamoudi et al. [14] proposed a fusion model incorporating ultrasound images and clinical data to diagnose PCOS, along with a basic categorization of the features used in their study. Despite these advances, current research exhibits several barriers: lack of comprehensive investigations into PCOS detection features and their interrelationships; insufficient exploration of the state of datasets used for detection; and lack of in-depth studies on the capabilities of existing intelligent PCOS detection tools.

In this paper, we propose an analytical framework for the current status of PCOS diagnostic research, which is a novel multi-stage approach designed to address these barriers. In the first stage, we conducted a rigorous literature analysis to identify the most relevant studies related to PCOS intelligent detection. In the second stage, based on the features mentioned in these selected studies and supplemented by diagnostic checklists provided by medical specialists, we constructed the comprehensive taxonomy of PCOS detection features to date. Additionally, we provided detailed annotations for each feature, including the acquisition methods and difficulty levels. In the third and fourth stages, based on the taxonomy, we analyzed the current status of datasets used for PCOS detection and conducted an in-depth evaluation of the capabilities of existing intelligent detection tools. Based on this research framework, we answer three key research questions (RQs):

- **RQ1:** What are the features used to detect PCOS so far, and what is their relevance? Through the systematic literature review, we identified 93 of the most relevant studies. Through the analysis of these studies, we constructed a comprehensive taxonomy of PCOS detection features, encompassing 110 features across eight categories.
- **RQ2:** What is the current status of datasets used for PCOS detection? We analyzed 12 publicly available datasets. These datasets collectively cover 58 features, with an overall coverage rate of 52% compared to our taxonomy. Additionally, we observed that none of the publicly available datasets are multi-modal, some have not been updated for years, and most lack clear license information.
- **RQ3:** What are the capabilities of current intelligent PCOS detection tools? Our analysis of 45 intelligent detection tools filtered from the relevant studies revealed that, while many perform well on test datasets, they still have the following limitations: high training costs, limited multi-modal processing capabilities, and lack of clinical validation.

In light of these findings, our research deepens the understanding of PCOS feature taxonomy and intelligent detection tools, offering valuable insights for future studies.

## 2. Background and Related work

This section introduces the background of PCOS detection and related work in three aspects:Machine Learning-based Detection Tools, Deep Learning-based Detection Tools, Reviews of PCOS detection.

### 2.1. Background of PCOS Detection

PCOS is a complex endocrine disorder with an unclear etiology, but it is widely believed to be associated with genetic factors, insulin resistance, and elevated levels of androgens in the body. Figure 1 shows normal ovaries and polycystic ovaries. The symptoms of PCOS are varied and may include irregular menstruation, infertility, hirsutism, acne, and weight gain. Additionally, many patients with PCOS also exhibit characteristics of metabolic syndrome, such as hypertension and dyslipidemia [8]. The detection of PCOS typically relies on the Rotterdam criteria, which require the presence of at least two of the following three conditions: oligo-ovulation or anovulation, clinical or biochemical signs of hyperandrogenism, and ultrasound findings of polycystic ovaries [9]. There have been previous studies aimed at applying intelligent algorithms to PCOS detection[12, 13, 14].

**Figure 1:**
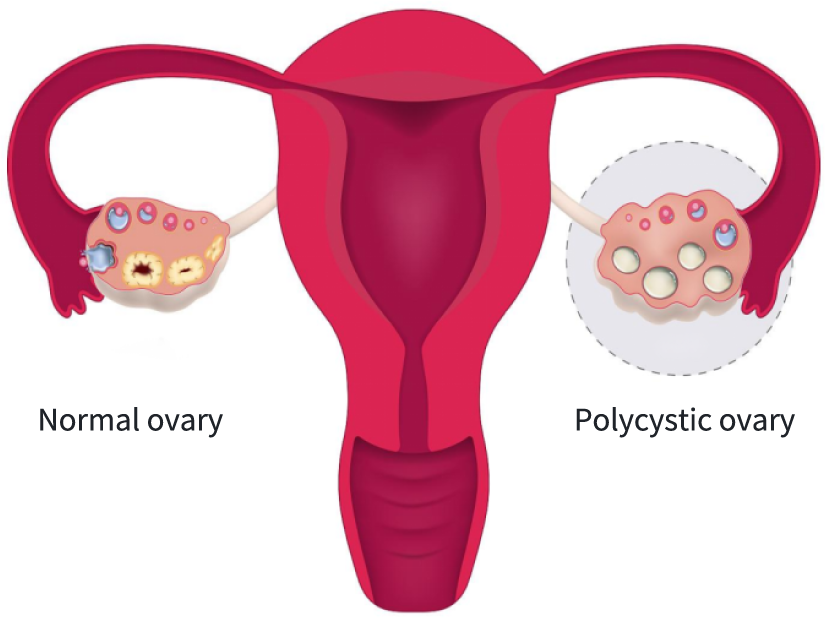
Polycystic Ovary and Normal Ovary Illustration

Figure 2 shows the stakeholders and overall process related to the use of intelligent algorithms in PCOS detection. The stakeholders include patients seeking treatment, researchers employing artificial intelligence for diagnosis, and physicians making comprehensive diagnostic decisions. In the diagnostic process, when patients initially visit the hospital, doctors conduct preliminary diagnoses through medical history collection and physical examinations. The collected physical information is then processed using artificial intelligence, which grades features such as the severity of hair loss, acne, and hirsutism. Based on these preliminary findings, doctors may request further examinations, including ultrasound and biochemical tests. All data obtained during the examinations are recorded in electronic medical records, and researchers utilize this data through AI-assisted diagnostic tools for intelligent PCOS diagnosis. Physicians integrate all information to make comprehensive diagnostic decisions. Finally, the results of the diagnosis are communicated back to the patients. It is worth noting that current PCOS intelligent detection tools have not yet been applied in clinical practice, as discussed in Section 4.3.3.

**Figure 2:**
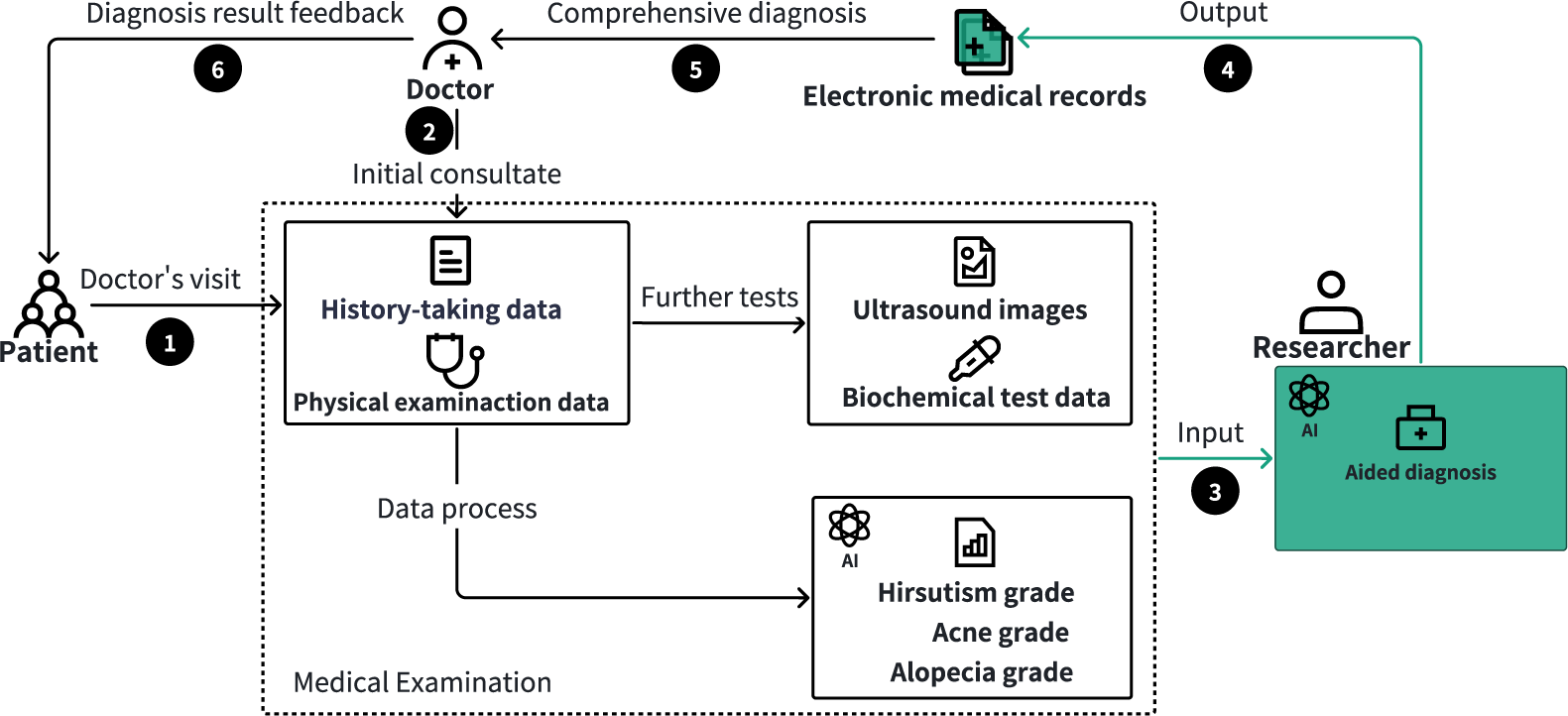
Stakeholders and processes related to PCOS intelligent detection

Our focus in this process is particularly on the stage of intelligent PCOS detection, where feature information is input into AI for assisted detection, as highlighted in the light green part of the diagram. This paper provides a systematic literature review of the PCOS intelligent detection field and establishes a taxonomy of PCOS features involved in the detection process.

### 2.2. Machine Learning-based Detection Tools

In PCOS intelligent detection, machine learning techniques are primarily applied in classification and feature selection.

The main machine learning classification techniques used for PCOS detection include support vector machines (SVM), decision trees, and ensemble methods. Specifically, Deshpandei et al. [15] employed the SVM algorithm to classify clinical, biochemical, and imaging data for PCOS diagnosis. Chauhan et al. [16] compared several machine learning algorithms and found that decision tree classifiers (DT) were among the most accurate models for predicting PCOS. Based on this, they developed a mobile app to help users predict PCOS in its early stages. Denny et al. [12] proposed a method with a minimal yet effective set of clinical and metabolic parameters. Zhang et al. [17] used a stacking classifier model based on k-nearest neighbors, random forests, and extreme gradient boosting to analyze Raman spectra of follicular fluid and plasma for PCOS screening.

In addition to classification algorithms, researchers have also delved into feature selection in PCOS detection to optimize performance. For instance, Zigarelli et al. [18] used principal component analysis (PCA) to extract features from highly correlated variables, achieving dimensionality reduction. Subha et al. [19] employed three traditional methods (i.e., correlation ranking, chi-square test (Chi2), and recursive feature elimination) along with two swarm intelligence methods (i.e., particle swarm optimization and firefly algorithm) for feature selection to identify significant PCOS features. Syed et al. [20] combined RF feature importance and highest correlation (HC) methods to select 10 features and used AdaBoost to achieve the highest detection accuracy for PCOS.

In summary, machine learning techniques in PCOS detection primarily focus on classification and feature selection, by combining classification algorithms and feature selection methods, these techniques improve diagnostic accuracy and efficiency. In contrast, this paper reviews and compiles the features used by various tools, constructs a taxonomy comprising 110 detection features, and provides a deeper analysis of the capabilities and limitations of existing machine learning-based detection tools.

### 2.3. Deep Learning-based Detection Tools

Deep learning techniques have shown significant potential in PCOS detection, particularly in image and data processing. Convolutional neural networks (CNN) have been widely applied to PCOS ultrasound image feature extraction and automatic detection due to their outstanding performance in handling image data. Specifically, Srivastav et al.[21] used a pre-trained VGG16 model to extract key features with CNN for PCOS detection. CNNs have also been applied to scleral image processing, where Lv et al. [22] proposed a deep learning-based method for screening PCOS through scleral image analysis.

In addition to image processing, deep learning techniques are also suitable for handling clinical, biochemical, and other types of one-dimensional data for automatic PCOS diagnosis. For example, Abouhawwash et al. [23] processed a dataset containing 39 features using deep learning models, including CNN, multilayer perceptron (MLP), recurrent neural networks (RNN), and bidirectional long short-term memory networks (Bi-LSTM). Additionally, Kumar [24] introduced a novel chaotic red deer optimization algorithm (CRDODL-BDADC) combined with big data analysis techniques based on deep learning, applying it to PCOS detection. Thomas et al. [25] proposed a deep learning method using CNN and the Adam optimization algorithm to classify PCOS-related terms in ultrasound text reports, enabling PCOS prediction and diagnosis. By analyzing textual data, this study highlights the versatility of deep learning applications in PCOS detection.

In summary, deep learning techniques in PCOS detection mainly focus on processing and analyzing images and One-dimensional data, enhancing the accuracy and efficiency of PCOS detection. Based on this, this paper reviewed deep learning-based PCOS detection tools and conducted a comprehensive evaluation of their capabilities and limitations.

### 2.4. Existing Reviews of PCOS Detection

In addition to the above-mentioned studies on PCOS detection based on intelligent algorithms, there are also reviews focusing on PCOS detection. Specifically, Modi [26] conducted a comprehensive analysis of AI applications in predicting lifestyle diseases, including PCOS, highlighting models, symptoms and risk factors, dataset selection, and research challenges that contribute significantly to healthcare research. In comparison, Suha [27] focused on computer-aided approaches for PCOS detection, achieving five main objectives: analyzing research aims and history, evaluating data sources and algorithms, summarizing research gaps, and proposing future directions. Barrera et al. [28] systematically reviewed studies applying machine learning and artificial intelligence (AI/ML) techniques for diagnosing and classifying PCOS, showcasing high diagnostic accuracy, and recommending future efforts to enhance standardization and methodological improvements for better clinical applications.

Previous reviews have mainly focused on models used in prior research. In contrast, this paper systematically reviews relevant literature, to construct a comprehensive taxonomy of PCOS detection features. Based on the taxonomy, we analyze the capabilities and limitations of current diagnostic tools and assess the state of datasets.

## 3. Research Design

In this section, we begin by presenting three research questions along with the motivations behind them. Subsequently, we introduce the overall framework of the empirical study and provide a detailed description of each module and step within it.

### 3.1. Research Questions (RQs)

- RQ1: What are the features used to detect PCOS so far, and what is their relevance? PCOS detection research is currently fragmented, lacking a comprehensive understanding of the features used. A thorough taxonomy of PCOS detection features would help us better understand the existing features and their relationships. The motivation behind establishing such a taxonomy is to provide a structured overview, facilitating clearer communication between researchers and clinicians and identifying the most relevant features for effective diagnosis. Currently, no such taxonomy exists, making this a critical gap in the field.
- RQ2: What is the current status of datasets used for PCOS detection? A major obstacle in PCOS detection research is the unclear status of the datasets. There is a lack of in-depth analysis regarding the current datasets, particularly concerning their openness, scale, licensing, and maintenance status. The purpose of conducting a comprehensive analysis of the current datasets is to identify their limitations, assess their availability, and guide future data collection efforts to improve the quality and accessibility of datasets for detection research.
- RQ3: What are the capabilities of current intelligent PCOS detection tools? While PCOS intelligent detection tools have demonstrated certain detection performance in their respective datasets, their capabilities are not fully investigated. The aim of understanding these tools’ capabilities is to evaluate their strengths and weaknesses, thus enabling the selection of the most appropriate tools and guiding the development of more accurate and efficient detection methods.

### 3.2. Research Framework

To address the three research questions outlined above, we propose an analytical framework for the current status of PCOS diagnostic research, based on the taxonomy construction method employed by Ladisa et al.[29]. As illustrated in Figure 3, the framework consists of four modules.

**Figure 3:**
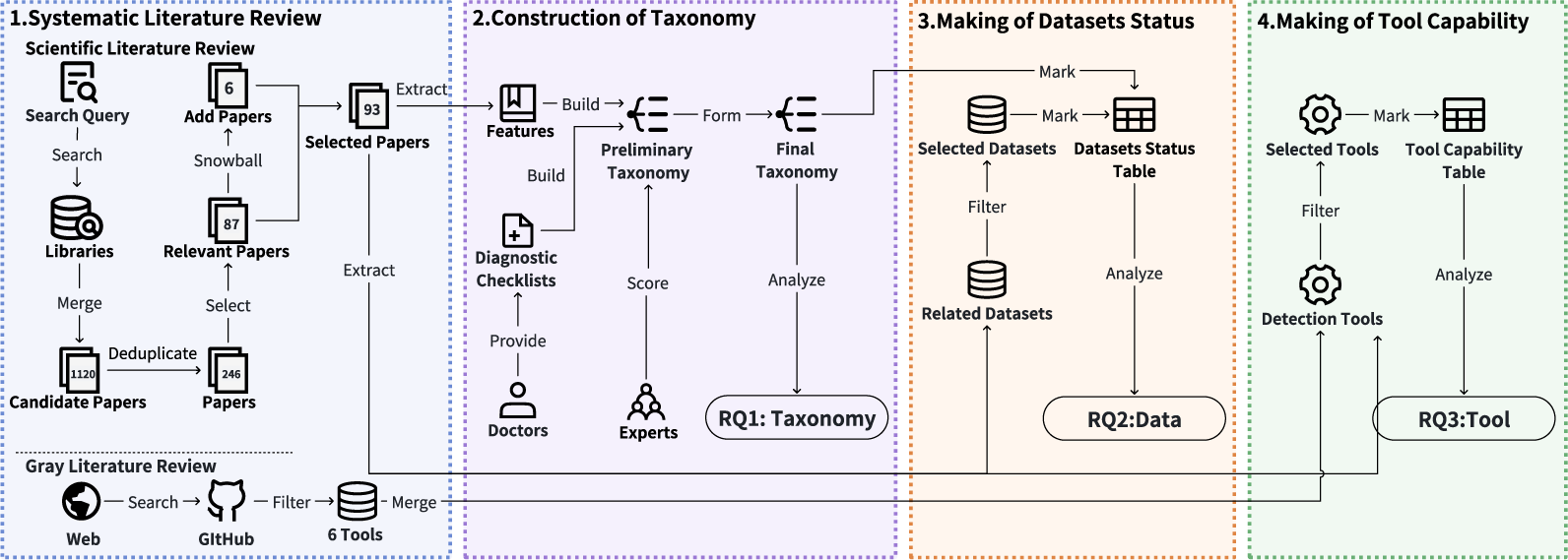
The framework of our study

Fitst, in the systematic literature review module, we obtained candidate literature through our constructed search query, then filtered it and conducted snowballing to identify the final selection of literature most relevant to intelligent PCOS detection. Subsequently, we extracted relevant information from these papers and explored grey literature. Second, in the taxonomy construction module, based on the features we collected and the diagnostic checklist provided by experts, we constructed a preliminary taxonomy for PCOS detection features. We then conducted a survey inviting experts to evaluate and provide feedback on the preliminary taxonomy’s rationality, completeness, and practical utility. Feedback indicated a high level of endorsement for the taxonomy’s rationality, completeness, and practical utility. Finally, based on this feedback, we finalized the preliminary taxonomy and formed the final taxonomy for PCOS detection features, thereby addressing RQ1. Third, in datasets status making module and tools capability making module, based on our taxonomy, we filtered the eligible datasets and representative tools, and assessed the current status of datasets and the capabilities of existing tools used for detection, answering RQ2 and RQ3.

### 3.3. Systematic Literature Review

This section provides a systematic review of scientific literature and grey literature related to PCOS intelligent detection, corresponding to the first module in Figure 3. To achieve this, we followed a structured approach for literature searching, which included multiple stages [30, 31].

#### 3.3.1. Scientific Literature Review

Scientific literature review consists of three stages: candidate literature searching, literature filtering and information extraction.

##### Candidate Literature Searching

The systematic literature review on PCOS detection research began by designing effective search queries [32, 33]. First, to ensure a comprehensive search, we conducted an exploratory search on Google Scholar using the keyword “PCOS detection.” This preliminary search found four relevant studies [19, 34, 35, 36], providing initial information on the topic and helping us identify relevant keywords. Subsequently, based on these initial studies, we carefully analyzed their keywords and created a search query string to retrieve articles most relevant to PCOS intelligent detection. Let the keyword sets be *K*_1_*, K*_2_*, K*_3_*,…, K_n_*, the keywords within each set are synonymous or closely related terms, representing similar concepts or phrases used in the context of PCOS detection, where:

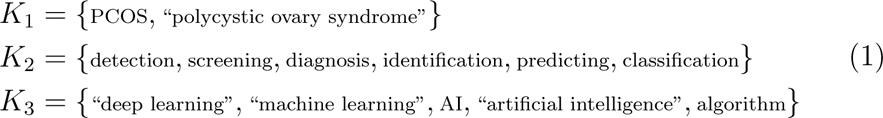

The search query string *Q* can be expressed as:

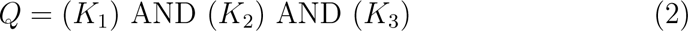

Using this carefully designed search query, we conducted extensive literature searches across six authoritative databases:IEEE Xplore, Scopus, Google Scholar, ACM Digital Library, PubMed and Web of Science. The search was limited to titles, abstracts, and author keywords, including only English publications, covering journals, conference proceedings, and book chapters. For each database *D ∈ {*Google Scholar, Scopus, IEEE Xplore*,…}*, we define the number of papers retrieved from each database as *n*(*D*). The total number of papers retrieved is:

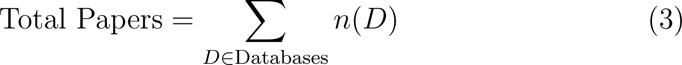

The specific numbers are:

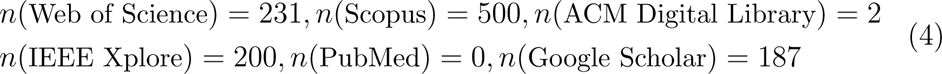

Therefore:

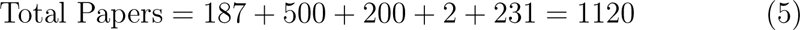

It should be noted that results from the PubMed medical database were excluded because they were not highly relevant to intelligent PCOS detection. This search process was completed by April 7, 2024.

##### Literature Filtering

The literature filtering process was conducted in four steps to identify the most relevant papers on PCOS intelligent detection. First, since there are duplicate entries, we carefully removed duplicate entries from the initial 1,120 candidate papers. After this step, 386 papers proceeded to the next round of screening. Second, to ensure that the selected literature was directly related to PCOS intelligent detection, we filtered the papers further. We examined the metadata of each paper, such as the type of publication, title, and abstract, to filter out papers unrelated to the topic. After this step, 246 papers remained. Third, to ensure sufficient information for our review, we obtained the full texts of all articles and focused on the introduction, methods, and results sections. Articles had to describe a PCOS detection tool and dataset, be over four pages, and in English; those not meeting these criteria were excluded. To maintain accuracy, all articles were independently reviewed by at least two authors, with final inclusion based on consensus, resulting in 87 relevant articles. Finally, to ensure comprehensive coverage, we applied a bidirectional snowball strategy, tracing forward and backward references from the 87 selected articles. Only references directly cited by or citing the selected papers were included, with a one-layer depth to maintain focus. This process added six papers, resulting in a final selection of 93 relevant papers for our study on PCOS intelligent detection.

Figure 4 shows the annual publication trend of the 93 selected papers. As depicted by the blue trend line, the number of publications has been steadily increasing. This trend indicates that research in the field of intelligent PCOS detection has been growing, reflecting the growing attention from both academia and the research community. This growth can likely be attributed to technological advancements and the increasing demand for early diagnosis and precise treatment of PCOS. It is worth noting that the number of selected papers for 2024 has decreased, as our paper collection cutoff date was April 7, 2024, and only papers published before that date were included.

**Figure 4:**
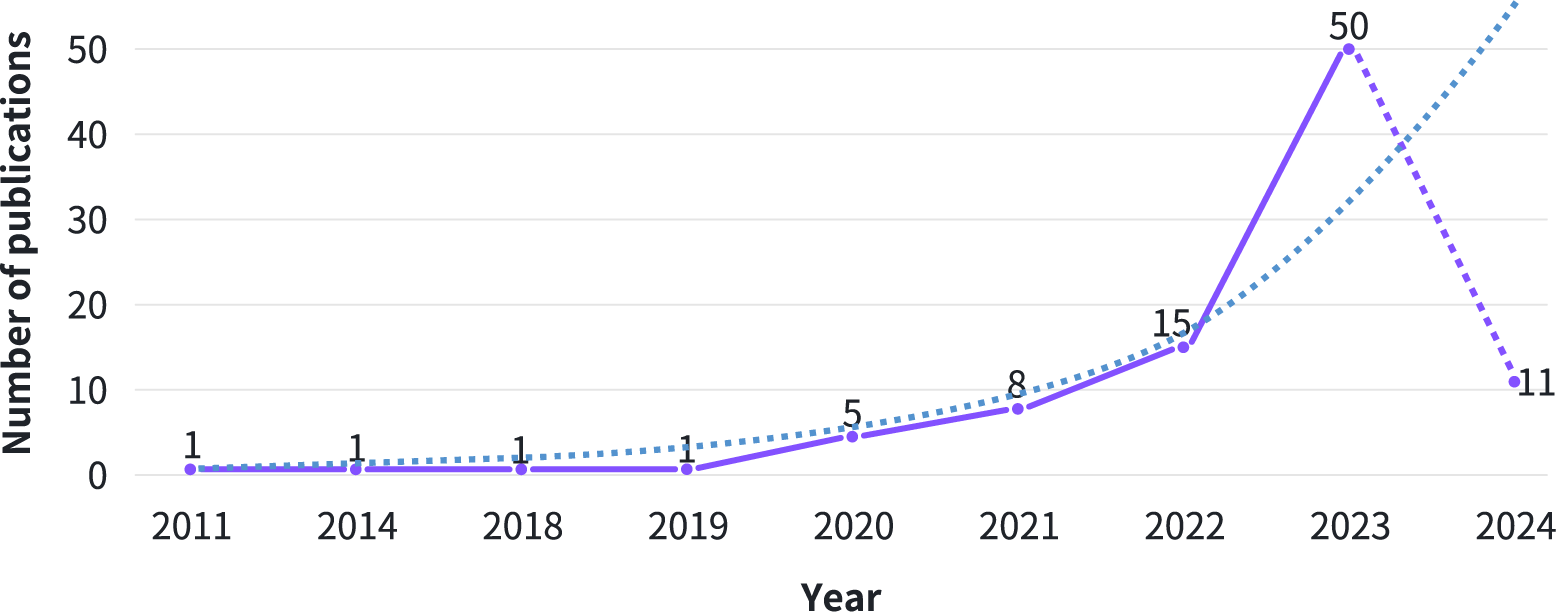
Annual Trends in PCOS Intelligent Detection Research Publications

##### Information Extraction

At this stage, we designed a structured extraction table to extract information from the papers. The extracted information includes the features used for detecting PCOS, the techniques employed, and the datasets utilized. Each paper was independently annotated by at least two authors to ensure data accuracy and consistency. Any discrepancies were resolved through discussion and review, resulting in consistent and high-quality data. This data serves as a critical foundation for answering the research questions.

#### 3.3.2. Grey Literature Exploration

In addition to scientific literature, we explored grey literature sources, such as blog posts, technical forums, and GitHub, the largest open source software website [29]. This stage aimed to comprehensively cover intelligent PCOS detection tools, enhancing the depth of our research. We regularly reviewed various repositories and blogs, using the same search query in Google as for scientific literature. The preliminary query revealed that grey literature related to PCOS detection was mainly concentrated on the open-source platform GitHub. We applied the same criteria used for scientific literature to filter these sources and used a snowball sampling strategy to expand the literature pool, ensuring thorough coverage. From the initial grey literature search, we identified 24 relevant entries. After further filtering, projects without a star were excluded. Ultimately, we identified 6 valuable open-source projects.

### 3.4. Taxonomy Construction

This section outlines the approach used to develop a taxonomy for PCOS detection features, as illustrated in the second module of Figure 3. Although previous studies have used feature selection methods to identify the features most relevant to PCOS [34],[36],[37], a systematic feature taxonomy has not yet been established. The goal was to construct a comprehensive taxonomy. The construction process consists of two consecutive steps: developing the preliminary taxonomy and expert survey.

#### 3.4.1. Preliminary Taxonomy Construction

We adopted a multi-step approach to constructing the preliminary taxonomy, which included three main steps: processing features extracted from the literature, systematically classifying these features, and annotating features. First, to eliminate redundancy, we refined the data on PCOS detection features extracted in Section 3.3.1 by merging similar features. Two types of similarity were considered: textual similarity, where features had identical definitions, synonymous terms, or referred to the same characteristic; and conceptual similarity, where features used different terminology but shared clinical or pathological significance. For merged features, we recorded both the original features and the unified name, detailed information is available on our website [38]. Each feature was manually verified, with two authors independently reviewing each category based on textual descriptions, examples, and attributions. Disagreements were resolved through the arbitration of a third author.

Second, building on the collected information, we classified the PCOS diagnostic features. We began by consulting with a hospital physician (third author) to obtain diagnostic checklists used in clinical practice. We matched the collected features against this checklists and, in consultation with the doctors, categorize these features into the corresponding categories on the checklists. Additionally, based on the physician’s extensive clinical experience, features used in actual diagnosis but not found in prior research were added to the taxonomy. These newly added features from clinical practice are marked with an asterisk to distinguish them from those documented in the literature.

Third, we annotated the acquisition methods and difficulty levels for each feature. We consulted with a specialized clinician(fourth author) to first identify how each feature is obtained during hospital visits. Then, based on the standard methods of acquisition in clinical testing, resource consumption, and the complexity of detection, we categorized the PCOS detection features into three levels:

- Level 1 (Easily Acquirable):These features are acquired through non-invasive and routine methods without requiring complex detection techniques or equipment. They typically involve self-reported information, simple physical measurements, or standard physical examinations.
- Level 2 (Moderately Acquirable): These features require specialized tests or equipment, often involving complex measurements or lab tests. While not highly invasive, the acquisition process may be time-consuming or require trained medical personnel to perform.
- Level 3 (Difficult to Acquire):These features necessitate highly complex detection methods or specialized equipment, potentially involving invasive procedures or advanced laboratory techniques, and are not part of routine testing. The acquisition process may be technically challenging and costly. In addition, some characteristics involve subjective assessments, and the accuracy of the assessment may be affected by individual self-reporting bias or different assessment criteria.

Through these steps, we constructed a preliminary taxonomy for PCOS detection features.

#### 3.4.2. Expert Survey

Following the development of the preliminary taxonomy, we conducted an expert survey to seek evaluation from industry professionals, including medical experts and researchers. The survey consisted of two parts: an online survey and expert interviews.

The online survey aimed to assess (a) the taxonomy’s rationality, completeness, and practical utility; and (b) the reasonableness of the annotations for acquisition methods and difficulty levels. It included 13 questions with specific evaluation objectives. Six questions used a Likert scale from 1 (low) to 5 (high) to assess various dimensions of the taxonomy [39]. Two questions were binary (yes/no), evaluating the use of the taxonomy and AI tools in PCOS detection. Additionally, four questions were used to collect demographic information such as age, work experience, and professional background of the participants. One question verified whether participants had read the taxonomy documents and background materials before rating, ensuring data quality. The online survey was conducted on the Wjx.cn platform from August 7 to August 21, 2024, and collected 87 responses. Anonymity was ensured through privacy measures, with no personal identification information collected. Detailed information about the survey can be found on our website [38].

Table 1 shows detailed demographic information of the 87 participating experts. This information includes age, years of professional experience, occupational background, familiarity with the concept and implications of PCOS. Results showed that 80% of the experts had over six years of experience, with 31 having more than 16 years. More than 95% of the experts are senior doctors and nurses, and 81% of the experts rated their understanding of PCOS as 4 or 5 points, whose expertise in disease diagnosis provides a solid guarantee for the validity of the taxonomy.

**Table 1:** Demographic Information of Experts.

| Demographic Information | Measure | Number | Percentage |
| --- | --- | --- | --- |
| Age | 18-25 | 6 | 7% |
|  | 26-30 | 15 | 17% |
|  | 31-40 | 30 | 34% |
|  | 41-50 | 26 | 30% |
|  | 51-60 | 10 | 12% |
| Years of work experience | 0-2 | 8 | 9% |
|  | 3-5 | 9 | 10% |
|  | 6-10 | 21 | 24% |
|  | 11-15 | 18 | 21% |
|  | Over 16 | 31 | 36% |
| Occupation | Medical professionals | 83 | 95% |
|  | Researcher | 1 | 1% |
|  | Other | 3 | 4% |
| Familiarity with PCOS | 5 (Familiar) | 30 | 35% |
|  | 4 | 41 | 47% |
|  | 3 | 11 | 13% |
|  | 2 | 3 | 3% |
|  | 1 (Unfamiliar) | 2 | 2% |

The expert interviews aimed to assess the practical utility, relevance, and applicability of the taxonomy in real-world clinical and research settings. One open-ended question aims to gather experts’ thoughts on the taxonomy and their insights on how it could improve clinical workflows and integrate with existing diagnostic tools.The question posed was: “In your clinical practice or research, how do you view integrating our PCOS taxonomy into existing diagnostic workflows or AI tools? Do you have any suggestions for adapting the taxonomy to different healthcare settings or detection tools?” The interviews, held between November 10 and 30, 2024, involved four experts. All data was anonymized, and privacy measures ensured confidentiality. Insights from these interviews were qualitatively analyzed to assess the taxonomy’s potential for clinical adoption.

The expert survey feedback recognized the preliminary taxonomy’s rationality, completeness, and practical utility, and leading to the adoption of the preliminary taxonomy as the final taxonomy.

### 3.5. Analysis of Datasets

In order to comprehensively understand the current status of PCOS detection datasets, we conducted a filtering and analysis of datasets relevant to PCOS detection.

#### 3.5.1. Filtering

We selected datasets from the 93 PCOS detection-related articles, focusing on whether these datasets meet the task’s relevance requirements. To this end, we established two core criteria:

- **Criterion #1 (Presence of PCOS Detection Instances):** The dataset must clearly include feature data for PCOS detection, ensuring researchers can extract instances for analysis and modeling.
- **Criterion #2 (Accessibility of the Dataset):** The dataset must be documented in the relevant publications and clearly indicate its availability to ensure practical support for research.

Applying these criteria, we selected 36 datasets for PCOS detection from the literature, numbered with the prefix “D” based on the review order.

#### 3.5.2. Analysis

We systematically analyzed the current status of these datasets across seven aspects: openness, instance types, scale, usage count, license information, maintenance status, and coverage.

First, to assess openness, we reviewed the availability of each dataset, categorizing them into open access, restricted access, and not publicly available, and documented access links and conditions for the accessible datasets. Second, we examined the “Methods” sections of relevant literature to classify the types of data present in each dataset, noting whether they comprised image data, numerical data, or other types. Similarly, to understand licensing terms, we reviewed the “Methods” sections of papers and visited dataset websites to confirm license types, identifying any restrictions on research and commercial use. The scale of each dataset was evaluated by counting the number of instances, and for datasets with the same scale, we further examined their similarities and differences by comparing the features contained in them. We analyzed usage frequency by counting how often each dataset was referenced in academic publications. Maintenance status was assessed by recording the last update date for each dataset to determine their current applicability in research. Finally, we evaluated the coverage of features included in the open source datasets compared to the 110 features we collected. Specifically, Column 6 of Table 2 lists the datasets that include this feature, indicating that the dataset “covers” the corresponding feature. This comprehensive approach provided a detailed understanding of the current landscape of PCOS datasets, highlighting areas for improvement in data availability and utility for research.

**Table 2:**
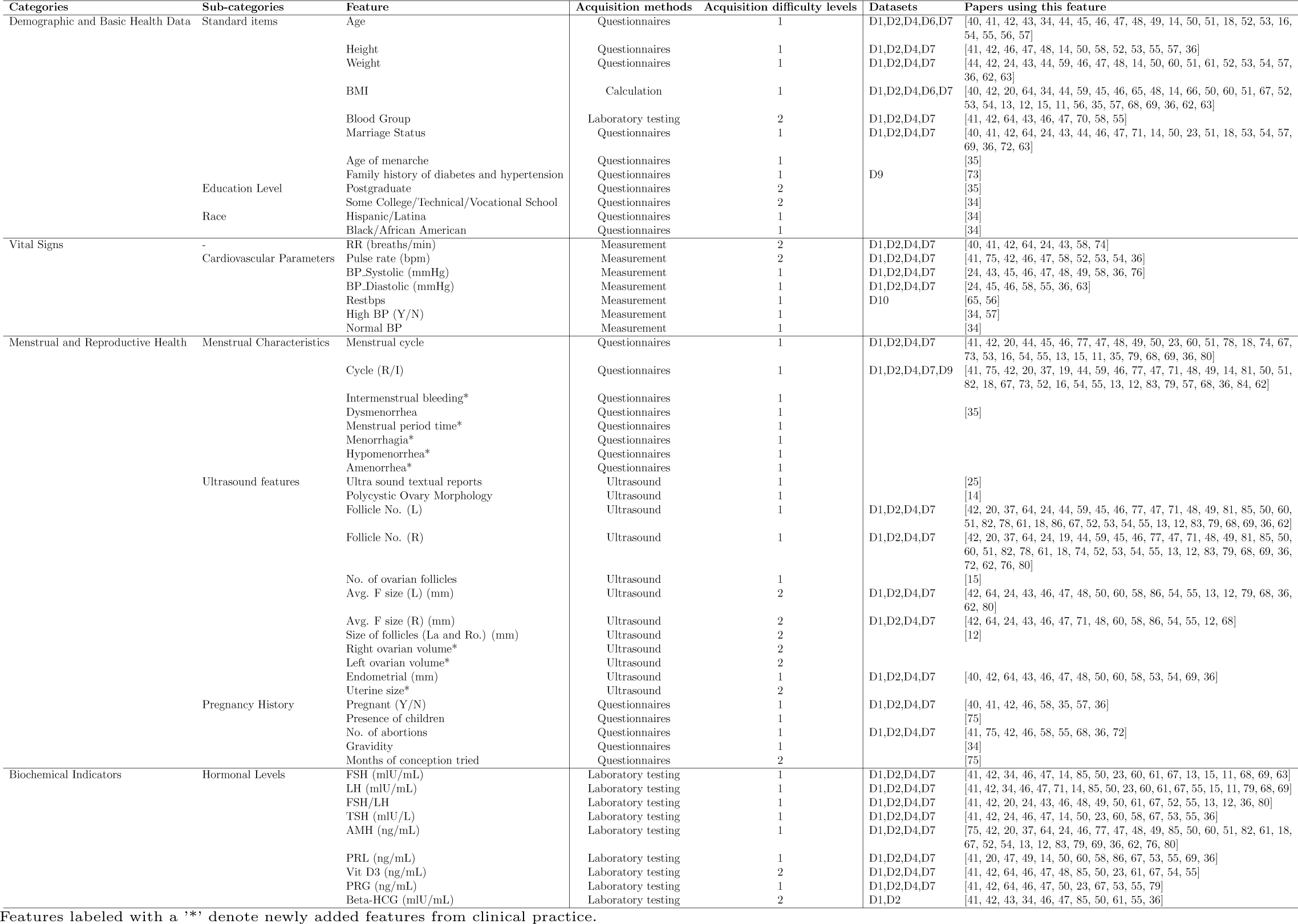
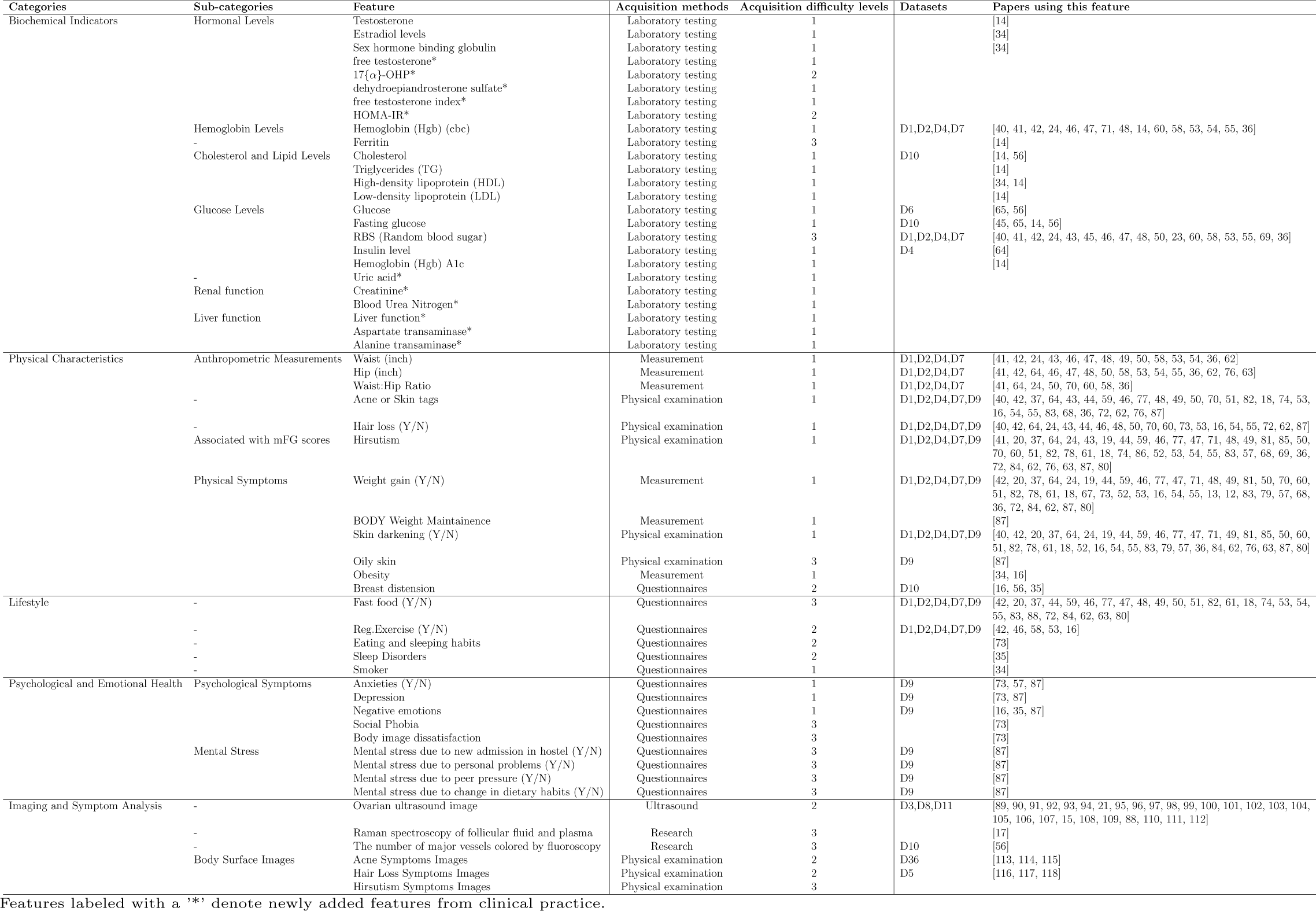
Taxonomy of PCOS detection features.

### 3.6. Analysis of Intelligent Detection Tools

Based on the 93 selected articles (see Section 3.3), we filtered out the representative tools according to established criteria and conducted an in-depth analysis of these intelligent PCOS detection tools.

#### 3.6.1. Filtering

We established the following criteria to ensure targeted analysis. Only tools meeting these criteria were selected:

- **Criterion #1 (Applicability to PCOS Detection):** To ensure that the tool is directly related to the PCOS detection task. The tool description should include terms such as “identification,” “detection,” or “classification,” and the output must be binary (PCOS or non-PCOS).
- **Criterion #2 (Automation):** Given the drawbacks of manual detection, such as inconsistent standards and inefficiency, we focused on tools with automated functions.
- **Criterion #3 (Industry Validation):** To ensure reliability, the tool must have been compared in other studies or its associated research must be indexed by SCIE.
- **Criterion #4 (Performance Metrics):** To facilitate valid comparison of tool performance in PCOS detection. The tool should record performance metrics such as accuracy, precision, and recall and disclose them transparently.

Following these criteria, we selected 39 eligible tools from the 93 articles, supplemented by six open-source PCOS detection tools from grey literature, resulting in a total of 45 representative tools. Since most tools lacked specific names, we assigned identifiers using the prefix “T” based on the review order.

#### 3.6.2. Analysis

We conducted a comprehensive analysis of these tools, covering aspects such as employed techniques, input data types, performance metrics, tool comparison and availability status. It should be noted that we did not run these tools ourselves; instead, we relied on the reported capabilities from their performance on the respective datasets.

First, we reviewed the techniques employed by each tool by examining the “Methods” sections of relevant papers. We extracted details about classification and feature selection algorithms, prioritizing combinations that achieved the highest accuracy as noted in the “Results” sections. These techniques were recorded using abbreviations, and we counted their usage frequency to identify the most commonly used techniques across the tools. Second, we identified the types of input data processed by each tool by documenting information from the “Methods” section of each paper. This allowed us to categorize tools based on the data types they handle, such as image, numerical, or text data. Third, we thoroughly assessed the performance metrics of each tool, focusing on the indicators reported in the “Results” sections. We recorded the performance achieved by the representative techniques of each tool and compiled the types of performance metrics reported. From this, we selected the four most commonly used metrics—accuracy, precision, F1 score, and recall—as the evaluation criteria. Tools were then categorized based on the data types they processed, and their performance metrics were analyzed to identify which tools demonstrated the best performance across different data types. Lastly, we reviewed the comparisons of representative tools documented in the selected papers, counting the number of times each tool was compared with others. We also documented their availability status, providing access links for open-source tools. This comprehensive analysis allowed us to evaluate the tools’ capabilities and influence within the research community based on the recorded information.

## 4. Results

Building on the framework outlined in section 3.2, this section provides responses to the three research questions.

### 4.1. Taxonomy

For RQ1, we constructed the taxonomy of features for PCOS detection based on 93 selected studies using the method described in Section 3.4.

Table 2 shows our taxonomy and the specific features of each category along with our annotated information. The taxonomy includes 8 categories and 32 subcategories, encompassing a total of 110 features relevant to PCOS detection. Figure 5 visually illustrates our taxonomy. We present the detailed results in three aspects: the categories, the methods and difficulty levels of feature acquisition, and the results of the survey.

**Figure 5:**
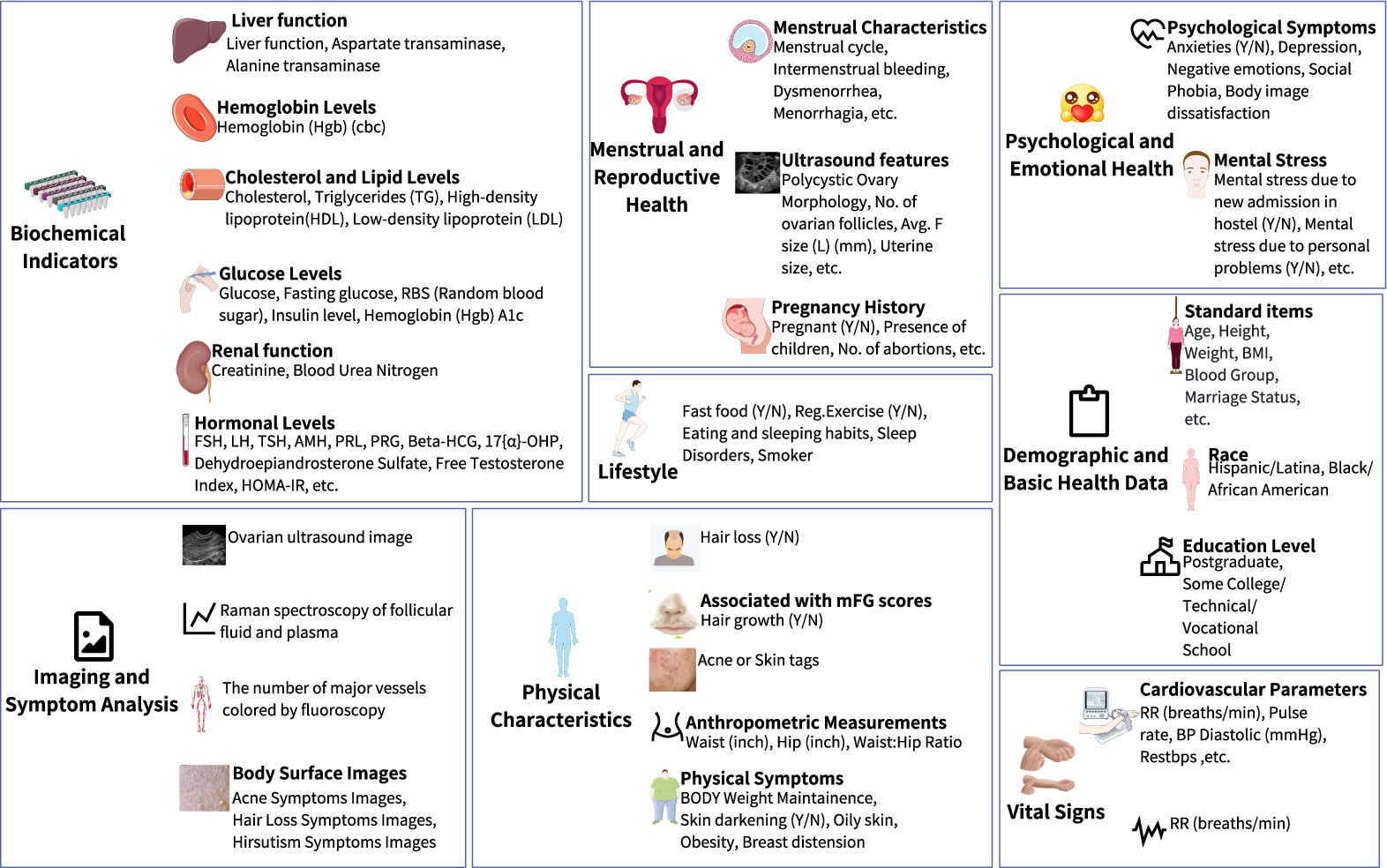
Features Illustrated in PCOS Detection Taxonomy

#### 4.1.1. Categories

Regarding specific categories, figure 6 shows the number of features contained in each category. The first category, “Demographic and Basic Health Data,” includes 3 subcategories and 12 features, mostly collected through questionnaires. BMI is the most commonly used feature in this category, appearing in 32 studies as an input for PCOS detection. The second category, “Vital Signs,” comprises 2 subcategories and 7 features, primarily obtained through medical measurements. Among these, systolic blood pressure (SBP) is cited in 10 studies, reflects the correlation between blood pressure and PCOS. The third category, “Menstrual and Reproductive Health,” is divided into 3 subcategories with 25 features, acquired via questionnaires and ultrasound. The Follicle No. (R) is frequently used, appearing in 40 studies, highlighting its role in evaluating ovarian function. The fourth category, “Biochemical Indicators,” is the largest, with 34 features spread across 8 subcategories, mainly derived from laboratory tests. Anti-Müllerian hormone (AMH) is used as a feature in 30 studies.

**Figure 6:**
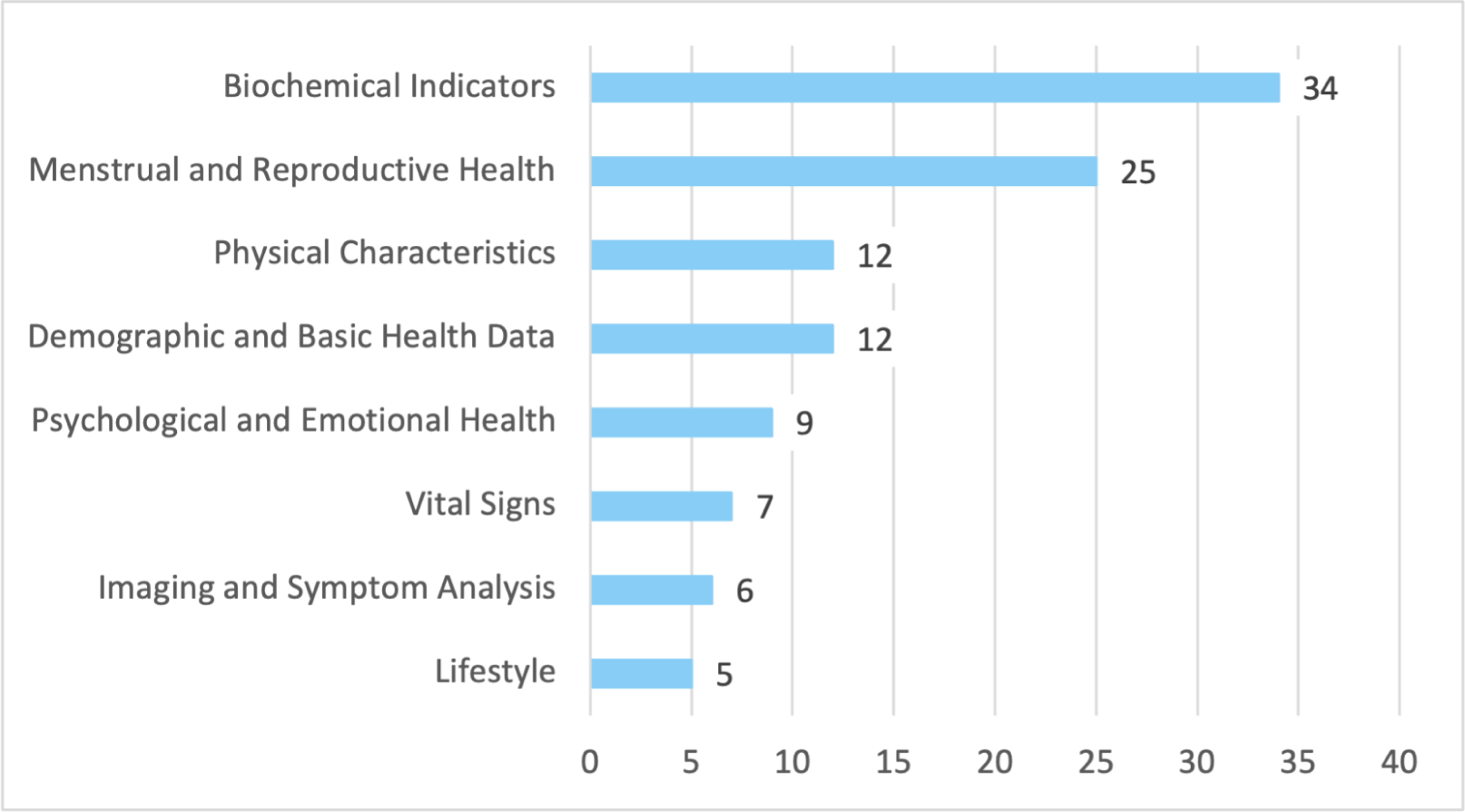
The number of features in each category

The fifth category, “Physical Characteristics,” comprises 12 features obtained through physical examination or medical measurements, with hirsutism and weight gain being cited in 43 studies, reflecting their significance as common PCOS symptoms. The sixth category, “Lifestyle,” includes 5 features collected via questionnaires. Fast food (Y/N) appeared in 26 studies, although features in this category are less frequently used, they offer valuable information about patient behavior. The seventh category, “Psychological and Emotional Health,” has 2 subcategories and 9 features, primarily gathered through questionnaires. The eighth category, “Imaging and Symptom Analysis,” includes 6 features such as image of acne, hair loss, and hirsutism, which are crucial in clinical diagnosis according to the *2023 international evidence-based guideline for the assessment and management of polycystic ovary syndrome* [3], although these surface image features have not been directly used to detect PCOS in existing intelligent tools.

In comparison, the biochemical indicators category contained the most features among the eight categories, while the lifestyle category contained the fewest features; hirsutism was the most frequently mentioned feature among the 110 features.

#### 4.1.2. Features acquisition methods and difficulty levels

Regarding feature acquisition methods, the taxonomy identifies seven methods of feature acquisition: questionnaires, calculations, laboratory tests, medical measurements, ultrasound, physical examination, and research data collection. Among these, 38 features are obtained through questionnaires, while BMI is the only feature derived from calculations.

In terms of acquisition difficulty levels, features are classified as easy, moderate, or difficult to obtain. The majority (74) are easy to acquire, with 23 moderately difficult and 13 difficult features. This classification aims to help researchers better understand and utilize these features, thereby selecting appropriate features and developing a more accurate and efficient detection tool while minimizing the cost and effort required for rapid diagnosis.

#### 4.1.3. Survey Results

Table 3 shows the results of the online survey. Table 4 shows the responses from the expert interviews. The taxonomy received strong support from industry experts.

**Table 3:** Feedback Results of Online Survey.

| # Feedback | # Valid Feedback | Structural Rationality | Classification Validity |
| --- | --- | --- | --- |
| 87 | 82 | 91% | 94% |
| Completeness | Practical Utility | Acquisition Methods Annotation | Difficulty Levels Annotation |
| 96% | 92% | 95% | 94% |

**Table 4:**
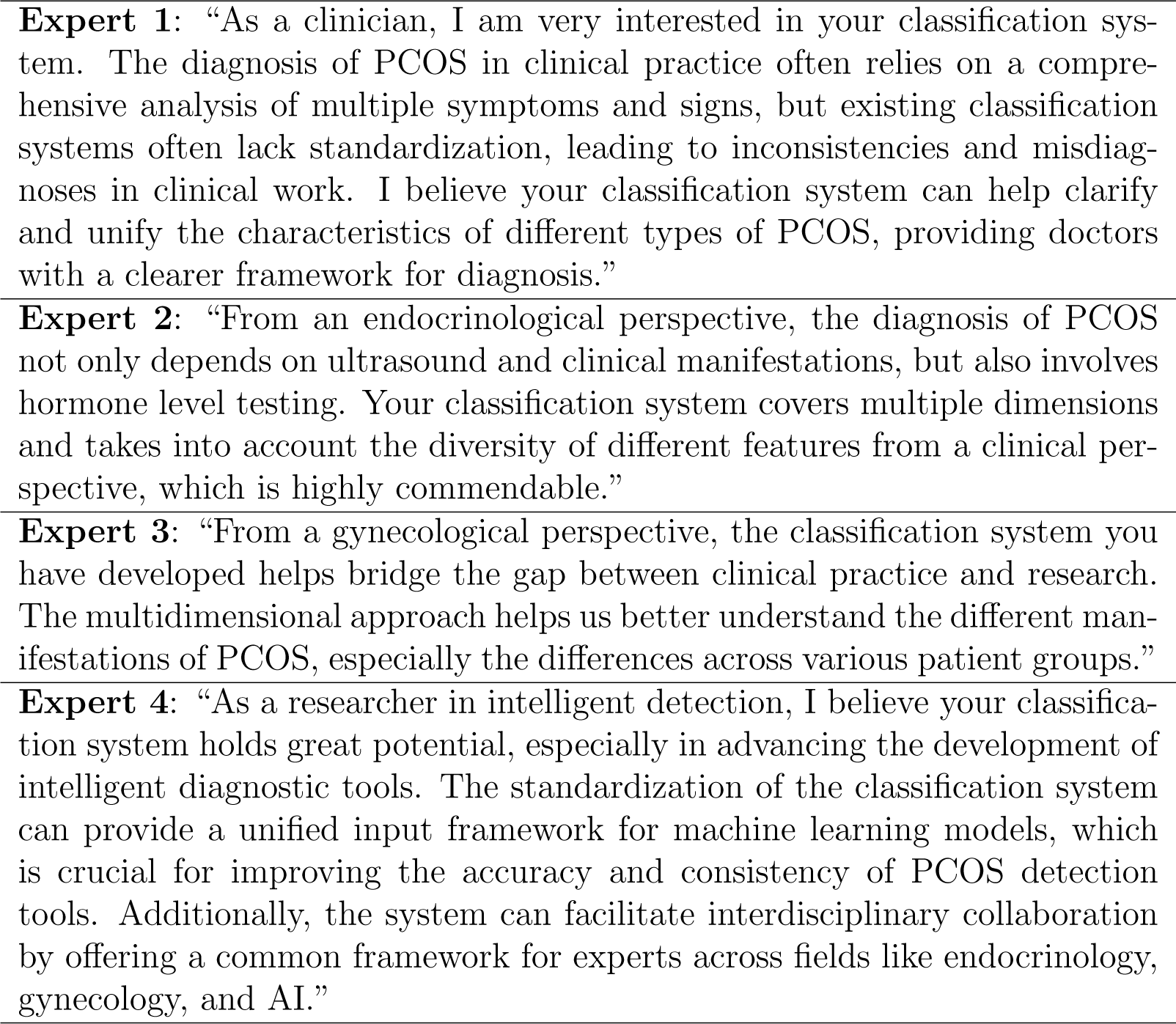
Responses of Expert Interview.

Specifically, in the online survey section, 91% of the 82 valid survey respondents rated the taxonomy as having structural rationality, classification validity, completeness, and practical utility, with high scores of 4 or 5. Over 94% of the participants Recognized our annotations regarding feature acquisition methods and difficulty levels. Additionally, the survey highlighted a gap in clinical practice, where only 48% of participants had used any PCOS-related taxonomy, and just 22% had experience with AI-based detection tools. This reveals the low popularity of artificial intelligence technology in the detection of PCOS in clinical practice, indicating a potential demand for promoting intelligent detection tools in the future. In the expert interview section, four experts expressed strong support for the taxonomy, highlighting its potential to standardize PCOS diagnosis across clinical, endocrinological, gynecological, and research perspectives, improving both clinical practice and the development of intelligent diagnostic tools.

**Answer to RQ1:** Through the review of 93 scientific publications, we have proposed a comprehensive taxonomy of PCOS detection features. This taxonomy is divided into eight categories, encompassing 110 detection features, and annotates the methods and difficulty levels of acquiring these features in clinical practice. The taxonomy was highly rated by 82 domain experts and researchers, including senior doctors and nurses.

### 4.2. Dataset Status

In analyzing RQ2, we aim to assess the current status of the selected datasets in seven aspects: openness, instance types, scale, usage count, license information, maintenance status, and coverage. Following the methodology outlined in Section 3.5, we identified 36 datasets containing instances of PCOS features.

As shown in Table 5, only the publicly accessible datasets and restricted datasets for PCOS detection are listed here; a complete list of all 36 datasets is available on our website [38]. In the following section, we present the specific analysis results.

**Table 5:** Datasets List for PCOS Detection.

| Dataset | Type | Scale | Usages | Openness | License | Year |
| --- | --- | --- | --- | --- | --- | --- |
| D36 | image | 1457 | 3 | Yes,[120] | - | 2019 |
| D5 | image | 4492 | 2 | Yes,[121] | - | 2021 |
| D11 | image | 1639 | 1 | Yes,[122] | - | 2022 |
| D3 | image | 3856 | 16 | Yes,[123] | - | 2022 |
| D8 | image | - | 1 | No,[119](Restricted) | - | - |
| D6 | numerical | 768 | 2 | Yes,[124] | CC0: Public Domain | 2016 |
| D9 | numerical | 119 | 1 | Yes,[125] | - | 2017 |
| D10 | numerical | 1273 | 2 | Yes,[126] | - | 2019 |
| D12 | numerical | 100 | 1 | Yes,[17] | - | 2020 |
| D2 | numerical | 541 | 40 | Yes,[127] | CC BY-NC-SA 4.0 | 2020 |
| D4 | numerical | 541 | 1 | Yes,[128] | - | 2020 |
| D7 | numerical | 541 | 2 | Yes,[129] | Data files © Original Authors | 2021 |
| D1 | numerical | 541 | 3 | Yes,[130] | - | 2022 |

#### 4.2.1. Openness

Regarding dataset openness, as shown in Figure 7a, among the 36 selected PCOS feature datasets, 23 are not publicly available, only 12 are open for access, and there is 1 restricted-access dataset. This indicates that most datasets used in research are not accessible to the public. The restricted dataset D8 [119] requires membership for access, and the specific number of instances it contains is unclear. Therefore, in subsequent statistics, any data involving instance counts will exclude this dataset. This quasi-open status effectively restricts researchers’ direct access to the data and impacts the usability and broad application of the datasets.

**Figure 7:**
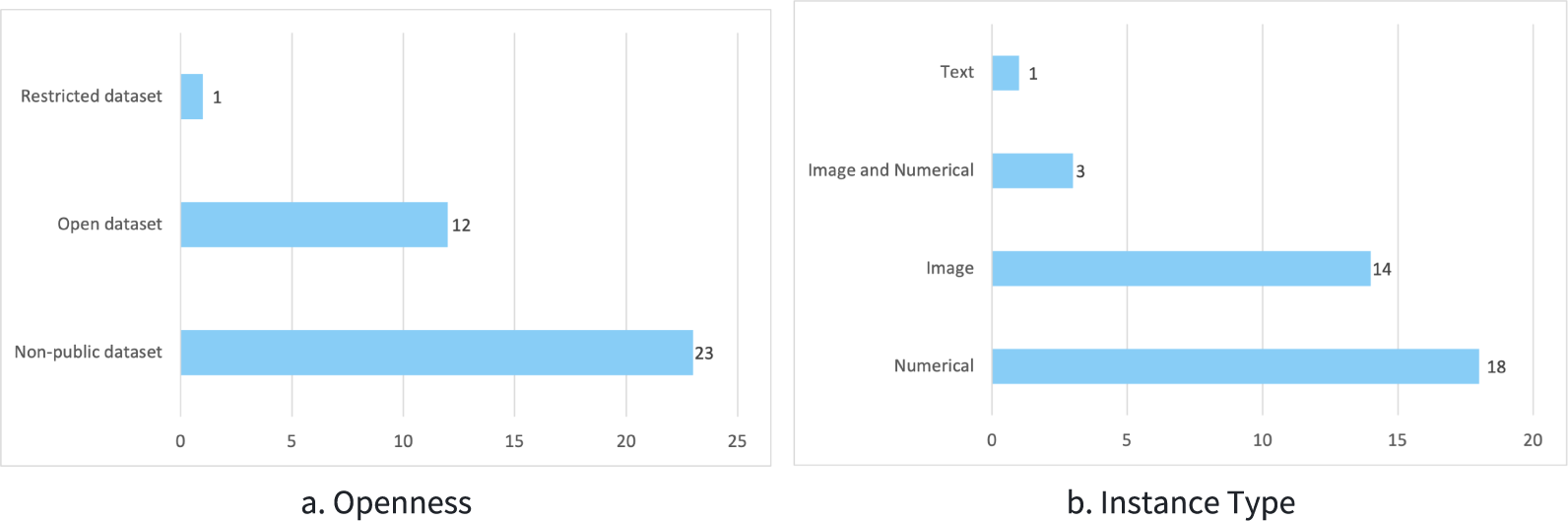
The Openness and Instance Type of the 36 Datasets

Insight: 67% of the PCOS datasets have restricted access, which significantly limits the research potential and hinders broader application.

#### 4.2.2. Instance Type

Concerning instance type, as shown in Figure 7b, the selected 36 datasets encompass image data, numerical data and text data. Specifically, 14 datasets consist of image data, 18 contain numerical data, 3 include both image and numerical data, and only 1 dataset is text-based. The lack of multimodal datasets (combining two or more instance types) hinders support for diverse diagnostic needs, limiting the generalization ability of existing tools.

Insight: The dominance of single-type datasets limits diagnostic versatility and tool adaptability in complex clinical scenarios.

#### 4.2.3. Scale

In terms of dataset scale, results indicate that image dataset D5 [113] contains the most instances (4492), significantly outpacing other image datasets. Among numerical datasets, D10 [126] with 1273 instances, is the largest. Notably, D1 [130], D2 [127], D4 [128], and D7 [129] share identical instance counts, and further examination indicates a high degree of overlap in features, suggesting these datasets may originate from the same primary dataset. Among them, D4[128] contains an additional insulin level feature compared to the other three datasets.

#### 4.2.4. Usage Count

D2 [127] and D3 [123] have the highest usage frequencies, referenced in 40 and 16 papers, respectively, containing 541 and 3856 instances. This suggests their widespread application in PCOS-related research, particularly D2 [127], which, due to its comprehensive numerical data, is frequently used for algorithm training and evaluation.

#### 4.2.5. License

Regarding license, among the 12 open-source datasets, only a few explicitly state their licensing information. Specifically, dataset D6 [124] operates under the CC0: Public Domain license, allowing unrestricted use without special permission, suitable for both public research and commercial applications. D2 [127] uses the CC BY-NC-SA 4.0 license, permitting researchers to share and modify the data for non-commercial purposes, provided they adhere to attribution, non-commercial use, and share-alike requirements [131]. This has contributed to D2’s prominence in academic citations. Additionally, dataset D7 [129] only states that the data file copyrights belong to the original authors, lacking further usage terms, which may impose limitations on public use.

Insight: Only three datasets have clear usage licenses, while most lack explicit licensing information, leading to uncertainty in their use and potentially preventing researchers from using them due to legal issues.

#### 4.2.6. Maintenance Status

Concerning maintenance status, among the 12 open-source datasets, the most recently updated include D11 [122], D3 [123], and D1 [130], all last updated in 2022. D6 [124] has not been updated since 2016, and D9 [125] last update was in 2017. Without sustained maintenance, affecting the applicability of models built on them, which is particularly problematic in fast-evolving fields like PCOS detection.

Insight: The lack of regular updates in datasets poses the risk of obsolescence, which can undermine the accuracy and reliability of models in PCOS detection research.

### 4.2.7. Coverage

Regarding dataset coverage, as shown in Table 2, these publicly available datasets collectively encompass 58 of the PCOS features identified in our classification system, yielding an overall coverage rate of 52% (58/110) compared to the known 110 features. Furthermore, a single dataset covers a maximum of 39 features and a minimum of 1 feature. Some datasets exhibit limited feature coverage, hindering comprehensive support for future developments and applications of PCOS detection tools. It is noteworthy that the coverage rate is merely a numerical value, and when combined with the performance metrics of detection tools discussed in Section 4.3, we found no significant correlation between the feature coverage of the datasets and the performance of the tools.

**Answer to RQ2:** Among the 36 PCOS datasets, 12 are publicly accessible, covering 58 features, which corresponds to an overall coverage rate of 52% compared to the known 110 features. At the same time, the current state of these datasets raises concerns due to the lack of multimodal datasets, many datasets have remained unupdated for years, and most lacking clear licensing information.

### 4.3. Capabilities of Intelligent Detection Tools

Addressing RQ3, this section aims to explore the capabilities of PCOS detection tools collected from scientific and grey literature. Following the process outlined in Section 3.6, we identified 45 representative PCOS intelligent detection tools.

Table 6 provides detailed information about these tools. In the following section, we present the analysis results from four aspects: the technologies used, the types of input data, the performance metrics of the tools, tool comparison and availability status.

**Table 6:**
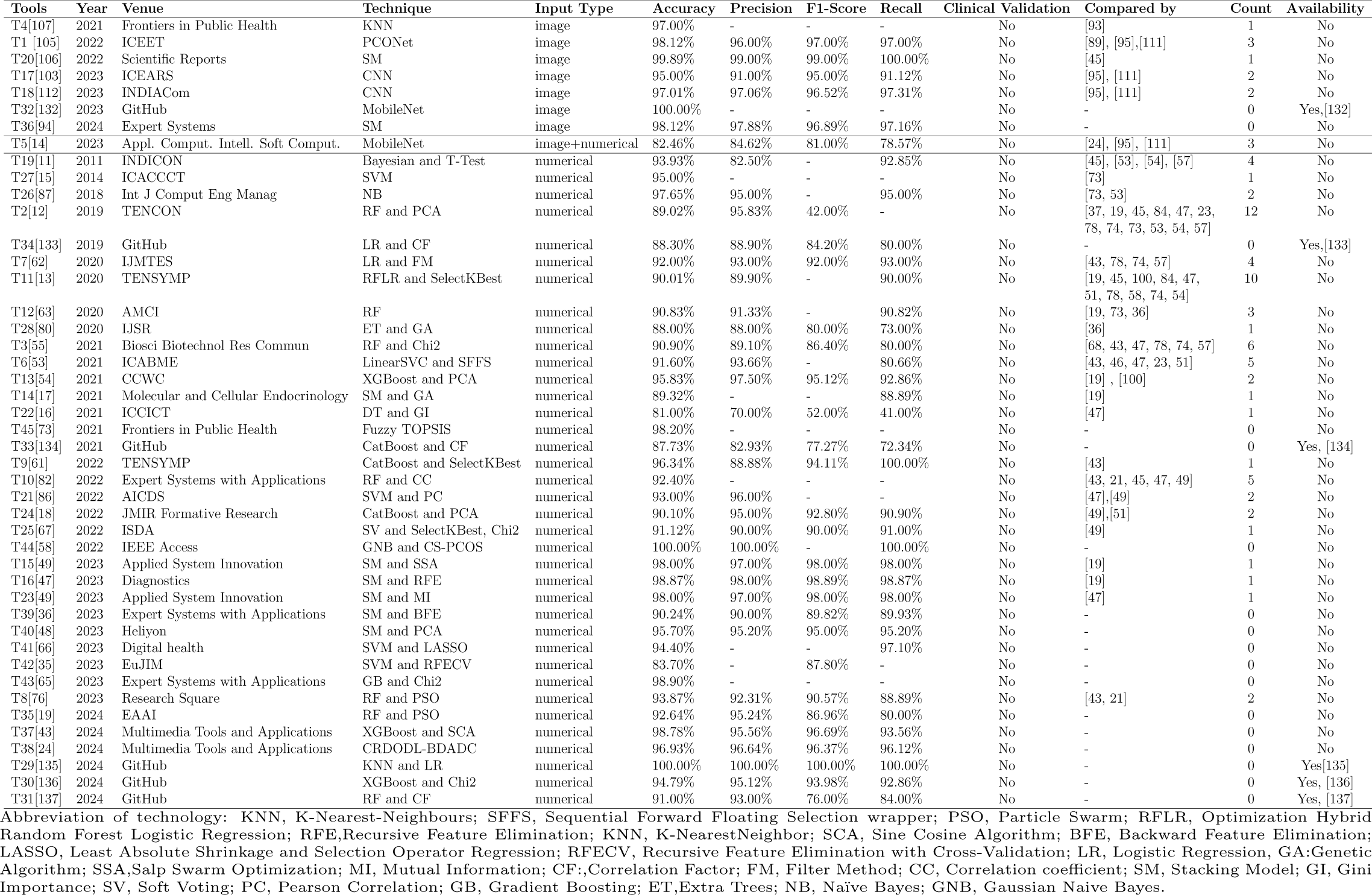
Tools List for PCOS Detection.

#### 4.3.1. Technology

In terms of technology employed, as shown in Figure 8a, we found that the vast majority (86%) utilize machine learning techniques, while only 13% employ deep learning. This indicates that machine learning remains the predominant approach for PCOS detection tools. Figure 8b shows the frequency of use of classification techniques, SM is the most widely used, with eight tools implementing this method. For feature selection methods, as shown in Figure 8c, PCA and Chi2 are the most frequently used, each employed by four tools. Tools using deep learning primarily employ CNN and MobileNet architectures for tasks involving ultrasound images or classification tasks that combine images with numerical data. The training of these ML and DL algorithms typically requires substantial computational resources and time, potentially leads to high training costs and may also extend the model development cycle.

**Figure 8:**
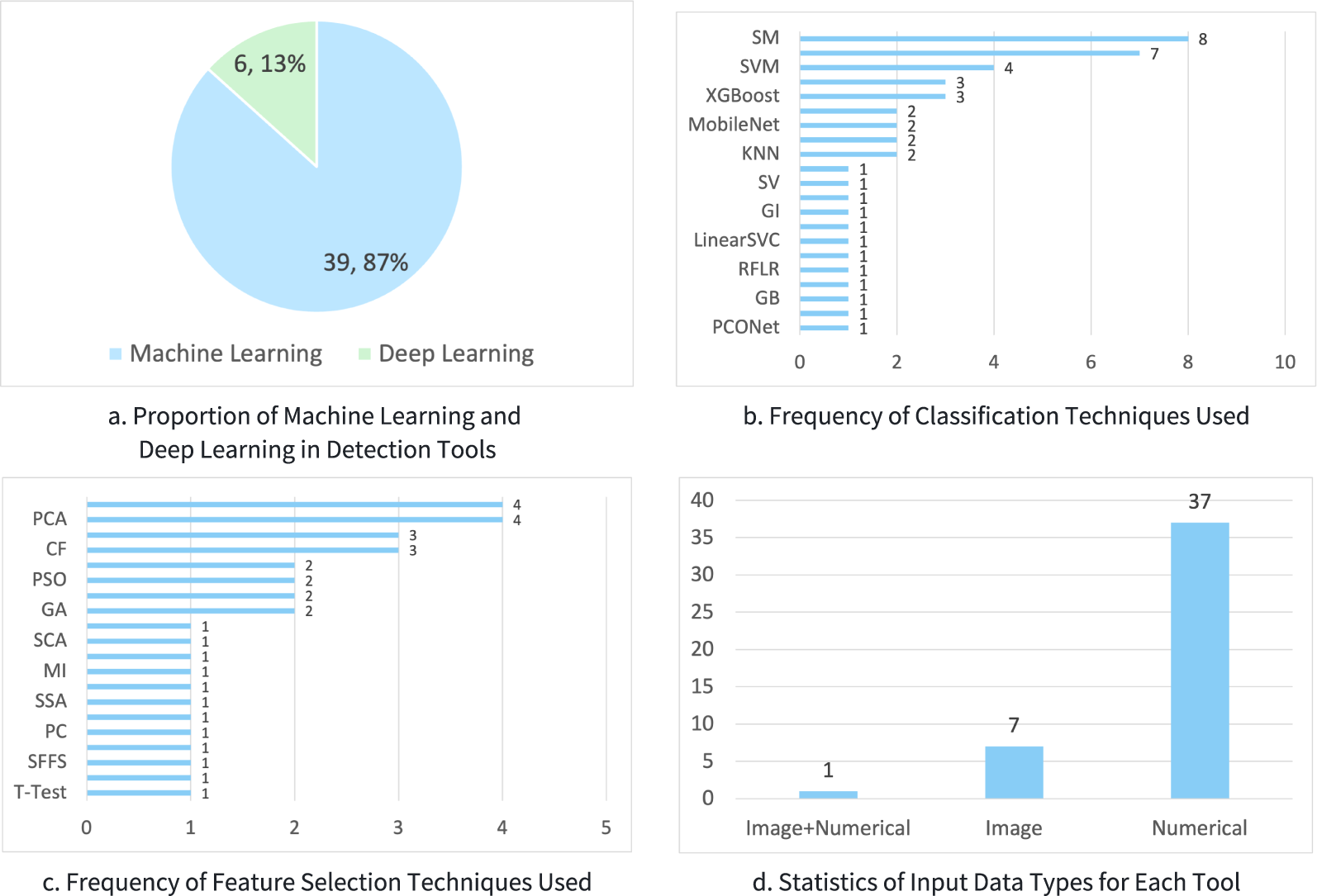
Statistics of Technologies and Input Data Types Used by Tools

Insight: The computational requirements of complex models may limit their popularization in resource-limited medical institutions, further hindering the widespread application of advanced detection technologies.

#### 4.3.2. Input Data Types

Regarding input data types, as shown in Figure 8d, among the 45 detection tools, 7 utilize image data, while 37 rely on numerical data. Only tool T5[14] combines image and numerical data as input. Only one tool uses multimodal data. The dominance of single data-type tools suggests that current PCOS detection tools may not fully capture the complexity of clinical cases. Insight: The limited integration of multimodal data in current tools underutilizes the potential for enhanced diagnostic accuracy.

#### 4.3.3. Performance Metrics

Performance metrics for various PCOS detection tools reveal significant variability, especially in their application of image and numerical data. Among the tools utilizing image data, T32[132] stands out with an impressive accuracy of 100%. Tool T20[106] also demonstrates remarkable performance, achieving precision and F1 scores of 99%, along with a recall rate of 100%. These results illustrate the potential of image data in PCOS detection, particularly when tools can effectively leverage complex visual information for high diagnostic precision. Similarly, tools using numerical data have also shown strong detection capabilities. Tools T44[58] and T29[135] achieved both 100% accuracy and precision, indicating that numerical data can also provide accurate PCOS detection, especially when combined with feature selection and classification algorithms. Moreover, T9[61] reached a recall rate of 100%, highlighting its exceptional ability to correctly identify all positive cases. Additionally, Tool T2 [12] achieved an accuracy of 89.02% and precision of 95.83%, but its F1 score was significantly lower at 42.00%. This discrepancy between precision and F1 score suggests that while Tool T2 excels at identifying true positives (high precision), it struggles with capturing enough of the true positives in the overall dataset (low F1 score). This highlights a potential issue in balancing sensitivity and specificity, suggesting that the tool may miss a considerable number of positive cases despite its high precision. Furthermore, as shown in column 10 of Table 6, although many tools demonstrate promising results on test datasets, they lack validation in real-world clinical settings.

Insight: Some tools demonstrate high accuracy on test datasets, but real-world validation remains crucial for PCOS tools.

#### 4.3.4. Tool Comparison and Availability Status

In analyzing the comparison frequency and availability of tools, T2 [12] and T11 [13] stand out as they are frequently compared, being used by 12 and 10 other tools, respectively. This highlights their significant influence in the academic community. Interestingly, the tools that are most frequently compared are not open-source. Among all the tools, only those derived from grey literature are open-source, and these open-source tools have not been subject to comparison by other tools. This trend suggests that while open-source tools may have a lower influence in academic circles, they nonetheless contribute to expanding our understanding of intelligent detection tools.

Insight: The dominance of closed-source tools in academic comparisons limits transparency and broader adoption, while open-source tools, though underrepresented, offer key benefits for scalability and collaboration.

**Answer to RQ3:** The selected 45 representative PCOS detection tools are all based on machine learning and deep learning technologies and demonstrate good detection performance on their test datasets. The detection accuracy of some tools even reached 100%. However, these tools still have the following limitations: (1) they require a lot of computing resources and take a long time to train; (2) they are insufficient in multimodal data processing; and (3) they lack clinical validation.

## 5. Discussion

This section discusses three key aspects: takeaway, limitations of our study, and challenges for future research.

### 5.1. Takeaway

Our research has constructed a comprehensive taxonomy of PCOS detection features, offering significant insights for both academia and industry.

- *Standardization*: As Expert 1 mentioned in Table 4. Researchers and medical professionals will benefit from the taxonomy, which offers a standardized framework for PCOS detection features. The taxonomy eliminates ambiguities and facilitates clear communication between researchers and industry experts, ultimately enhancing their understanding of detection features.
- *Data Evaluation*: Data scientists will find valuable insights for future data collection and usage strategies. We conducted a detailed analysis of the datasets used for PCOS detection, evaluating aspects such as openness, size, maintenance status, and coverage. This process uncovered deficiencies in feature coverage and the availability of multimodal data, providing a clear direction for future dataset improvements.
- *Tool Evaluation*: Researchers in the field can utilize the results of the tool evaluation to refine existing tools or develop new methodologies that enhance diagnostic accuracy. By assessing the capabilities of existing PCOS detection tools, we identified their limitations, guiding the development of more efficient detection algorithms.
- *Interdisciplinary Collaboration*: As Expert 4 mentioned in Table 4. The taxonomy offers a common language and toolset for researchers across different fields, promoting interdisciplinary collaboration and supporting the integration of various diagnostic methods to enhance the comprehensiveness of PCOS detection. This fosters teamwork among medical researchers, data scientists, and healthcare practitioners, ultimately improving patient outcomes.

### 5.2. Limitations

This study has several limitations that may affect the generalizability and accuracy of the findings.

First, our taxonomy is based on PCOS detection features extracted from existing literature and publicly available datasets. Due to factors such as access limitations, language barriers, or database restrictions, some relevant studies or datasets may not have been included, potentially leading to selection bias. This may affect the completeness and accuracy of the taxonomy.

Second, while we incorporated expert feedback during the feature labeling process, the subjective nature of manual annotations could introduce some degree of inconsistency or inaccuracies in the feature classification. Additionally, due to time and resource constraints, we were unable to empirically validate all of the PCOS detection algorithms discussed in the literature, which means there could be discrepancies between reported and actual tool performance.

Third, our study relies on currently available public datasets, which may not fully represent the diversity and complexity of PCOS patients, especially in the absence of multimodal data, limiting the generalizability and applicability of our conclusions in different clinical settings.

Finally, as PCOS detection technologies and related algorithms rapidly evolve, our research, which is based on existing techniques and literature, may face challenges regarding the validity of the current taxonomy and conclusions as new technologies and methods emerge. Continuous updates will be necessary to maintain relevance and accuracy.

### 5.3. Challenges

This study identifies three major challenges in the detection of PCOS: the timeliness of taxonomy, limitations in existing datasets, and deficiencies in detection tools. Addressing these challenges is essential for effectively advancing future developments.

First, not all features included in the current taxonomy are used in actual clinical diagnosis. Our taxonomy encompasses 110 features, which, based on discussions with medical professionals, can be classified into three categories: features actively used in diagnosis, features considered with some uncertainty, and features primarily used in research contexts that have yet to be adopted clinically. Moreover, the lack of mechanisms for real-time updates limits the taxonomy’s adaptability to emerging diagnostic trends, biomarkers, and technologies. Future research should prioritize developing a dynamic and scalable taxonomy that integrates automated updates from new literature and datasets. Such advancements would ensure the taxonomy remains relevant, flexible, and capable of supporting evolving clinical needs, thereby enhancing its applicability in practical diagnostics.

Second, inadequacies in datasets and their associated limitations pose significant barriers. Many datasets suffer from restricted access, delayed updates, insufficient instances, or a lack of multimodal data. Many datasets are not publicly available or have restricted access, which limits their use. Although some datasets are widely used, the scarcity of datasets combining multiple data types hinders diagnostic accuracy and limits the generalization capabilities of detection tools. Furthermore, datasets that are publicly available often lack relevance to current clinical practices, further diminishing their utility. Addressing these issues requires constructing open, comprehensive, and diverse datasets that support the training of robust PCOS detection models, particularly those designed to integrate and process multimodal data effectively. This approach will better accommodate the growing complexity and diversity of real-world clinical diagnostic needs.

Finally, current intelligent detection tools face challenges related to computational complexity, resource demands, limited multimodal processing, and insufficient clinical validation. Tools primarily rely on machine learning and deep learning methods, such as CNNs, which, while powerful, demand substantial computational resources and time, increasing costs and lengthening development cycles. Additionally, most tools are constrained to single data types, which restricts their adaptability to complex clinical scenarios. Moreover, a lack of rigorous clinical validation limits confidence in their real-world diagnostic effectiveness and safety. Future research should emphasize developing efficient, multimodal detection models capable of processing diverse data types and ensuring robust clinical validation to establish their practicality in real healthcare settings.

In summary, addressing these challenges will lay a robust foundation for advancing PCOS detection. Priorities for future research include creating a dynamic taxonomy, constructing open and comprehensive multimodal datasets, and developing clinically validated, efficient detection tools. These efforts will collectively improve diagnostic accuracy, scalability, and real-world impact.

## 6. Conclusion

This study addresses critical barriers in PCOS detection research, specifically concerning the taxonomy of detection features, intelligent detection tools, and available datasets. We successfully constructed a comprehensive taxonomy of PCOS detection features to date based on 93 relevant papers. This taxonomy includes the types and sources of features, a list of relevant studies that utilize these features, and annotates the acquisition methods and difficulty levels for each. Based on this taxonomy, we conducted a review of existing intelligent detection tools, revealing their capabilities and limitations, and conducted an in-depth analysis of the datasets used for detection and evaluated the current status of the datasets. In future work, based on the development of a unified dataset, we will compare the capabilities of these tools and, building on this comparison, develop more efficient PCOS detection algorithms.

## Data Availability

All data produced in the present study are available upon reasonable request to the authors

## Notes

### Competing Interest Statement

The authors have declared no competing interest.

### Funding Statement

This work was partially supported by the NSFC International Collaboration and Exchange Progaram (No. W2412110), Science and Technology Program Project of Shenzhen(No. SZWD2021012), Natural Science Foundation of Top Talent of SZTU (grant no. GDRC202132), SZTU-Enterprise Cooperation Project(No. 20221061030002, and No. 20221064010094), Shenzhen Science and Technology Program (No. JCYJ20220818102215034).

